# Safety and immunogenicity of the Ad26.COV2.S COVID-19 vaccine candidate: interim results of a phase 1/2a, double-blind, randomized, placebo-controlled trial

**DOI:** 10.1101/2020.09.23.20199604

**Authors:** Jerald Sadoff, Mathieu Le Gars, Georgi Shukarev, Dirk Heerwegh, Carla Truyers, Anne Marit de Groot, Jeroen Stoop, Sarah Tete, Wim Van Damme, Isabel Leroux-Roels, Pieter-Jan Berghmans, Murray Kimmel, Pierre Van Damme, Jan de Hoon, Williams Smith, Kathryn E. Stephenson, Dan H. Barouch, Stephen C. De Rosa, Kristen W. Cohen, M. Juliana McElrath, Emmanuel Cormier, Gert Scheper, Jenny Hendriks, Frank Struyf, Macaya Douoguih, Johan Van Hoof, Hanneke Schuitemaker

## Abstract

**BACKGROUND:** The ongoing coronavirus disease (COVID)-19 pandemic caused by severe acute respiratory syndrome coronavirus 2 (SARS-CoV-2) might be controlled by an efficacious vaccine. Multiple vaccines are in development, but no efficacious vaccine is currently available.

**METHODS:** We designed a multi-center phase 1/2a randomized, double-blinded, placebo-controlled clinical study to assesses the safety, reactogenicity and immunogenicity of Ad26.COV2.S, a non-replicating adenovirus 26 based vector expressing the stabilized pre-fusion spike (S) protein of SARS-CoV-2. Ad26.COV2.S was administered at a dose level of 5×10^10^ or 1×10^11^ viral particles (vp) per vaccination, either as a single dose or as a two-dose schedule spaced by 56 days in healthy adults (18-55 years old; cohort 1a & 1b; n= 402 and healthy elderly ≥65 years old; cohort 3; n=394). Vaccine elicited S specific antibody levels were measured by ELISA and neutralizing titers were measured in a wild-type virus neutralization assay (wtVNA). CD4+ T-helper (Th)1 and Th2, and CD8+ immune responses were assessed by intracellular cytokine staining (ICS).

**RESULTS:** We here report interim analyses after the first dose of blinded safety data from cohorts 1a, 1b and 3 and group unblinded immunogenicity data from cohort 1a and 3. In cohorts 1 and 3 solicited local adverse events were observed in 58% and 27% of participants, respectively. Solicited systemic adverse events were reported in 64% and 36% of participants, respectively. Fevers occurred in both cohorts 1 and 3 in 19% (5% grade 3) and 4% (0% grade 3), respectively, were mostly mild or moderate, and resolved within 1 to 2 days after vaccination. The most frequent local adverse event (AE) was injection site pain and the most frequent solicited AEs were fatigue, headache and myalgia. After only a single dose, seroconversion rate in wtVNA (50% inhibitory concentration - IC50) at day 29 after immunization in cohort 1a already reached 92% with GMTs of 214 (95% CI: 177; 259) and 92% with GMTs of 243 (95% CI: 200; 295) for the 5×10^10^ and 1×10^11^vp dose levels, respectively. A similar immunogenicity profile was observed in the first 15 participants in cohort 3, where 100% seroconversion (6/6) (GMTs of 196 [95%CI: 69; 560]) and 83% seroconversion (5/6) (GMTs of 127 [95% CI: <58; 327]) were observed for the 5×10^10^ or 1×10^11^ vp dose level, respectively. Seroconversion for S antibodies as measured by ELISA (ELISA Units/mL) was observed in 99% of cohort 1a participants (GMTs of 528 [95% CI: 442; 630) and 695 (95% CI: 596; 810]), for the 5×10^10^ or 1×10^11^ vp dose level, respectively, and in 100% (6/6 for both dose levels) of cohort 3 with GMTs of 507 (95% CI: 181; 1418) and 248 (95% CI: 122; 506), respectively. On day 14 post immunization, Th1 cytokine producing S-specific CD4+ T cell responses were measured in 80% and 83% of a subset of participants in cohort 1a and 3, respectively, with no or very low Th2 responses, indicative of a Th1-skewed phenotype in both cohorts. CD8+ T cell responses were also robust in both cohort 1a and 3, for both dose levels.

**CONCLUSIONS:** The safety profile and immunogenicity after only a single dose are supportive for further clinical development of Ad26.COV2.S at a dose level of 5×10^10^ vp, as a potentially protective vaccine against COVID-19.

**Trial registration number:** NCT04436276

## INTRODUCTION

The first cases of coronavirus disease 2019 (COVID-19) caused by severe acute respiratory virus coronavirus 2 (SARS-CoV-2) were reported in Wuhan, China, in December 2019.^1, 2^ Since then, the virus has infected millions of people globally. Infection with SARS-CoV-2 can result in a range of clinical manifestations, varying from asymptomatic infection to severe acute respiratory distress and death. To halt this pandemic, and to stop the pressure on health care systems and the negative effects on the global economy, efficacious COVID-19 vaccines are urgently needed.

Several vaccine candidates are currently in different stages of development.^3–6^ We have developed Ad26.COV2.S, that is based on Janssen’s replication incompetent adenovirus serotype 26 (Ad26) vector, and a stabilized SARS-CoV-2 Spike (S) protein. This candidate was selected based on its immunogenicity and manufacturability profile.^7^ The Ad26 vector is also used for our Ebola vaccine, and RSV, HIV and Zika vaccine candidates and Ad26-based vaccines are generally well tolerated and highly immunogenic.^10^

We recently reported that a single dose of Ad26.COV2.S elicited strong immune responses in rhesus macaques, and upon challenge with SARS-CoV-2, none of the vaccinated animals had detectable viral load (VL) in bronchoalveolar lavage (BAL) and only one out of six had transient and low VL in the nose.^5^ Protective efficacy strongly correlated with the presence of virus neutralizing activity in serum of the animals. In addition, we demonstrated that Ad26.COV2.S provided protective immunity in a Syrian golden hamster severe disease model.^11^

These preclinical data supported the start of a Phase 1/2a randomized, double-blinded, placebo-controlled clinical study to assesses the safety, reactogenicity and immunogenicity of Ad26.COV2.S in one- or two-dose (8-week interval) schedules with 5×10^10^ or 1×10^11^ viral particles (vp) per vaccination in adults 18–55 or ≥65 years of age. Here we report interim blinded safety and reactogenicity data as well as group unblinded immunogenicity data obtained during the first 4 weeks after the first vaccination. Based on these results, we have initiated a Phase 3 study that will evaluate the efficacy of a single vaccination of 5×10^10^ vp of Ad26.COV2.S (Trial Number: NCT04505722).

## METHODS

### STUDY DESIGN AND PARTICIPANTS

We are performing a multicenter, randomized, double-blind, placebo-controlled Phase 1/2a trial to evaluate safety, reactogenicity and immunogenicity of Ad26.COV2.S at 5×10^10^ or 1×10^11^ vp, administered intramuscularly (IM) as single-dose or two-dose schedules, 8 weeks apart, in healthy adults 18–55 years of age (cohort 1a (target n=375; interim safety and immunogenicity results post dose 1 reported here) and cohort 1b (target n=25; interim safety results post dose 1 reported here)) and ≥65 years of age (cohort 3 (target n=375; interim post dose 1 safety on full cohort reported here, as well as interim immunogenicity of first 15 participants that were 65–75 years of age)). Enrollment of Cohort 2 will start later and will allow us to collect additional safety and immunogenicity data (see Supplementary Tabble 1 for details on cohort 2). Overview of the clinical trial design is given in Table 1 and in the flow chart in Figure 1.

**Table 1.** COV1001 study design.

| **Cohort 1a (Adults ≥18 to ≤55 years)** |  |  |  |
| --- | --- | --- | --- |
| **Group** | **N** | **Day 1 (Vaccination 1)** | **Day 57 (Vaccination 2)** |
| 1 | 75 | Ad26.COV2.S 5×10<sup>10</sup> vp | Ad26.COV2.S 5×10<sup>10</sup> vp |
| 2 | 75 | Ad26.COV2.S 5×10<sup>10</sup> vp | Placebo |
| 3 | 75 | Ad26.COV2.S 1×10<sup>11</sup> vp | Ad26.COV2.S 1×10<sup>11</sup> vp |
| 4 | 75 | Ad26.COV2.S 1×10<sup>11</sup> vp | Placebo |
| 5 | 75 | Placebo | Placebo |
| **Cohort 1b (Adults ≥18 to ≤55 years)** |  |  |  |
| **Group** | **N** | **Day 1 (Vaccination 1)** | **Day 57 (Vaccination 2)** |
| 1 | 5 | Ad26.COV2.S 5×10<sup>10</sup> vp | Ad26.COV2.S 5×10<sup>10</sup> vp |
| 2 | 5 | Ad26.COV2.S 5×10<sup>10</sup> vp | Placebo |
| 3 | 5 | Ad26.COV2.S 1×10<sup>11</sup> vp | Ad26.COV2.S 1×10<sup>11</sup> vp |
| 4 | 5 | Ad26.COV2.S 1×10<sup>11</sup> vp | Placebo |
| 5 | 5 | Placebo | Placebo |
| **Cohort 2a (Adults ≥18 to ≤55 years)** |  |  |  |
| **Group** | **N** | **Day 1 (Vaccination 1)** | **Day 57** |
| 1-4 | 120 | Ad26.COV2.S 5×10<sup>10</sup> vp | No vaccination |
| 5 | 15 | Placebo | No vaccination |
| **Cohort 2b (Adults ≥18 to ≤55 years)** |  |  |  |
| **Group** | **N** | **Day 1 (Vaccination 1)** | **Day 57 (Vaccination 2)** |
| 1-4 | 120 | Ad26.COV2.S 5×10<sup>10</sup> vp | Ad26.COV2.S 5×10<sup>10</sup> vp |
| 5 | 15 | Placebo | Placebo |
| **Cohort 3 (Adults ≥65 years)** |  |  |  |
| **Group** | **N** | **Day 1 (Vaccination 1)** | **Day 57 (Vaccination 2)** |
| 1 | 75 | Ad26.COV2.S 5×10<sup>10</sup> vp | Ad26.COV2.S 5×10<sup>10</sup> vp |
| 2 | 75 | Ad26.COV2.S 5×10<sup>10</sup> vp | Placebo |
| 3 | 75 | Ad26.COV2.S 1×10<sup>11</sup> vp | Ad26.COV2.S 1×10<sup>11</sup> vp |
| 4 | 75 | Ad26.COV2.S 1×10<sup>11</sup> vp | Placebo |
| 5 | 75 | Placebo | Placebo |
| **Total** | **1,045** |  |  |

**Figure 1:**
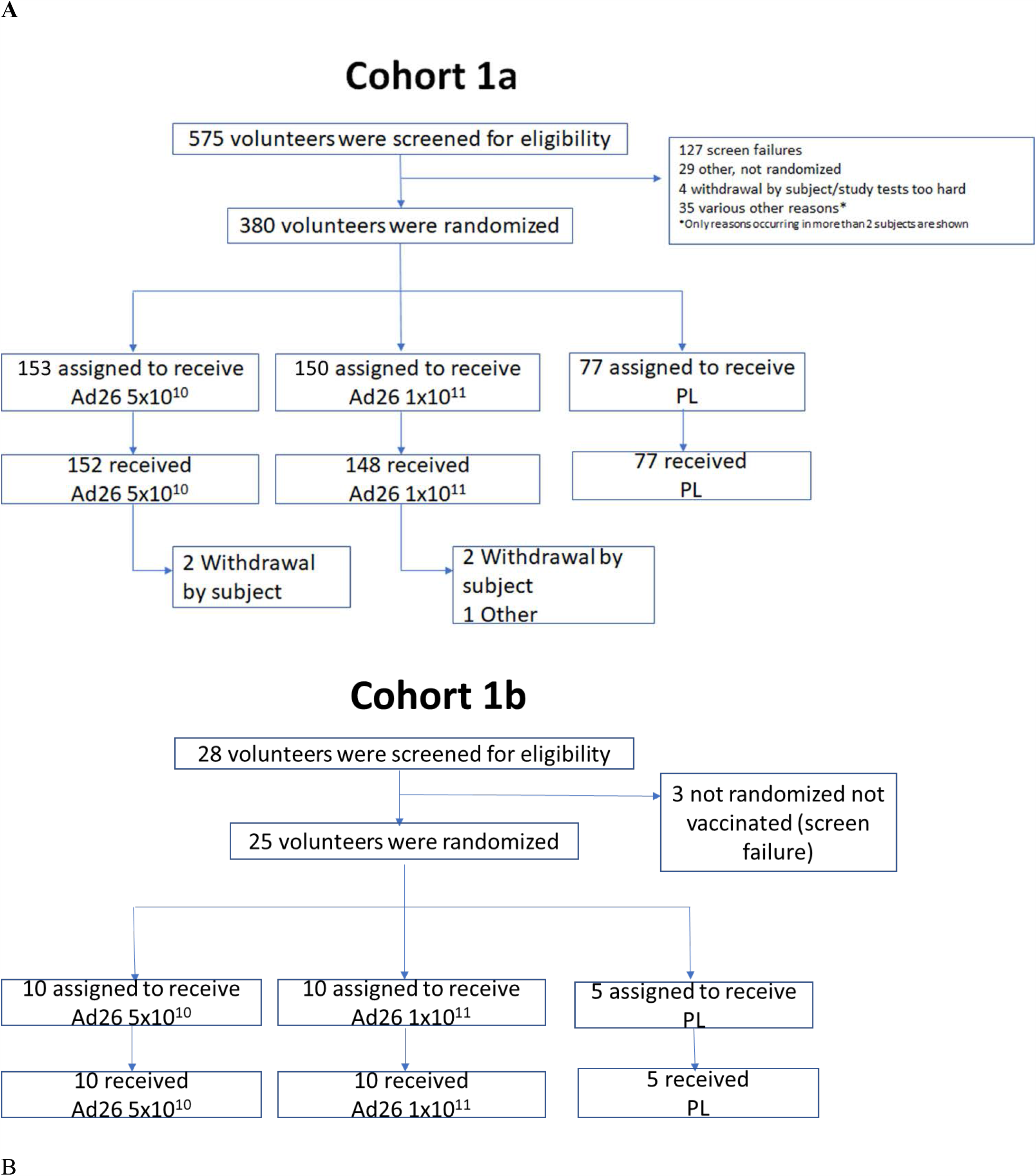
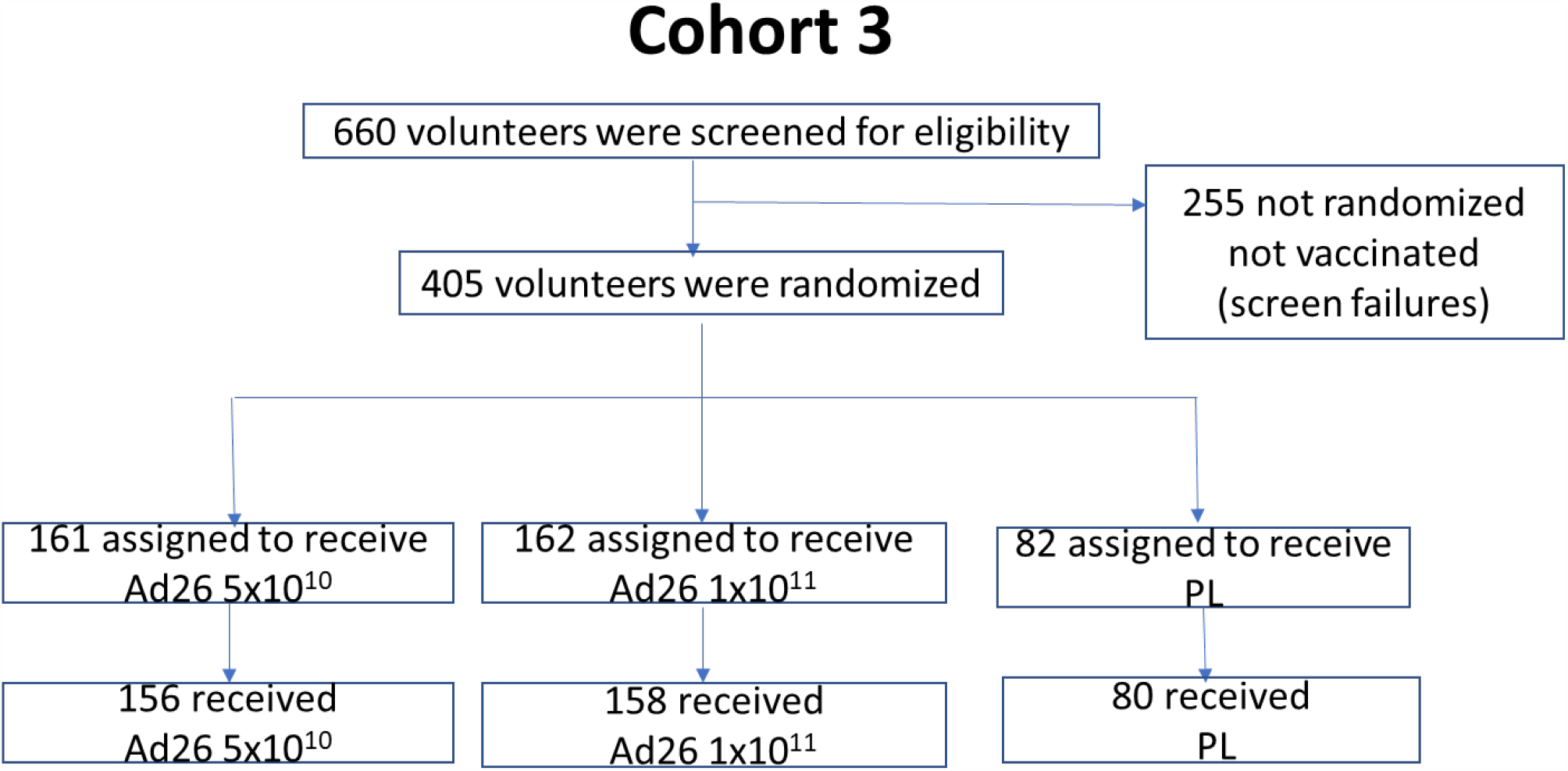
Consort Flow charts for cohort 1a, cohort 1b and cohort 3. **A** Participants were enrolled concurrently at Belgian and US sites. Participants were randomized in parallel in a 1:1:1:1:1 ratio to one of five vaccination groups to receive one or two IM injections of Ad26.COV2.S at two dose levels of either 5×10^10^ vp or 1×10^11^ vp, or placebo. For cohort 1 and 3, in the absence of clinically significant findings 24 hours after the first vaccination was administered to five sentinel participants (two per dose level and one placebo), another ten participants were vaccinated across all groups. Safety data up to Day 28 were then reviewed by an internal data review committee before the remaining participants were randomized.

Ad26.COV2.S is a recombinant, replication-incompetent Ad26 vector^12^ encoding the full length and stabilized SARS-CoV-2 S protein^7^ derived from the first clinical isolate of the Wuhan strain (Wuhan, 2019, whole genome sequence NC_045512). The study is performed at multiple clinical sites in Belgium and the United States. A list of inclusion and exclusion criteria is provided at ClinicalTrials.gov (Trial registration number: NCT04436276). All participants were screened for COVID-19 by the collection of nasal samples for PCR, which, if positive, would exclude them, and by locally available serological assays for detection of previous infection with SARS-CoV-2 with a maximum of 25 seropositive participants allowed between Cohort 1a and Cohort 3. The study was reviewed and approved by local ethics committees (Comité d’Ethique Hospitalo-Facultaire Sain-Luc, Université Catholique de Louvain on July 16, 2020) and institutional review boards (IRB) (approval by Advarra IRB on June 29 and July 10, 2020, for New Orleans Center for Clinical Research and Optimal Research sites, respectively). All participants provided written informed consent before enrollment.

### RANDOMIZATION AND BLINDING

We randomly assigned 405 eligible participants 18–55 years of age and 405 participants ≥65 years of age to receive one or two vaccinations with either a 5×10^10^ or 1×10^11^ vp dose level of vaccine, or placebo (1:1:1:1:1 per age group): 5×10^10^vp/5×10^10^vp; 5×10^10^vp/placebo; 1×10^11^vp /1×10^11^vp; 1×10^11^vp/placebo; or placebo/placebo for the first and second dose, respectively (Figure 1).

Randomization was done by an Interactive Web Response System (IWRS) and stratified by site using randomly permuted blocks. Participants and investigators remain blinded at individual participant level throughout the study. Vaccine and placebo were provided in masked identical syringes. Sponsor and statisticians were group-unblinded for the interim analysis when all participants completed the day 29 visit or discontinued earlier. Safety results are presented here in a blinded manner in order to prevent accidental unblinding.

### ENDPOINTS

Endpoints to support the primary objectives of safety and reactogenicity of each dose schedule were adverse events (AEs) for 28 days after each vaccination, local and systemic reactogenicity for 7 days after each vaccination, and serious adverse events (SAEs) throughout the study. AEs were graded according to FDA Guidance document “Toxicity Grading Scale for Healthy Adult and Adolescent Volunteers Enrolled in Preventive Vaccine Clinical Trials”. The secondary endpoint was humoral immune response to the S protein of SARS-CoV-2 as demonstrated by Spike-specific enzyme-linked immunosorbent assay (ELISA) and by neutralizing titers against wild type SARS-CoV-2 in a wtVNA, and cellular immune responses as measured by intracellular cytokine staining (ICS) after stimulation with S peptide pools.

### PROCEDURES

Participants received intramuscular (IM) injections of 5×10^10^ vp, 1×10^11^ vp Ad26.COV2.S or placebo (0.9% saline) in a 1 mL volume in the deltoid muscle at Day 1. Solicited AEs were collected on diary cards for 7 days post vaccination, unsolicited AEs for 28 days after vaccination and SAEs throughout the course of the study. Safety data are included up to the cut-off dates of 27 August 2020 (cohort 1) and 7 September 2020 (cohort 3). Baseline seropositivity was assessed by local clinical assays, and blood samples for serum chemistry, hematology, and for immune responses were and will be collected at several timepoints throughout the study; urine samples for pregnancy testing were collected before vaccination.

At baseline and on Day 29, Spike (S)-specific binding antibodies were measured by ELISA. Seropositivity was defined as a titer ≥50.3 EU/mL. SARS-CoV-2 serum neutralizing antibody titers were measured in a microneutralization wtVNA using the Victoria/1/2020 SARS-CoV-2 strain at Public Health England (PHE). Seropositivity in the wtVNA was defined as an IC_50_ titer ≥58. SARS-CoV-2 S-specific T-cell responses were measured at baseline and on Day 15 by ICS using two pools of S peptide pools of 15mers overlapping by 11 amino acids. A Th1 response was characterized by CD4+ T cells expressing IFN-γ and/or IL-2 and not IL-4, IL-5 and/or IL13; a Th2 response was characterized by CD4+ T cells expressing IL-4, IL-5 and/or IL-13 and CD40L by CD4+ T cells. All assays were conducted in a blinded fashion and are described in detail in the Appendix.

### STATISTICAL METHODS

This study was designed to assess safety and immunogenicity. Safety data were analyzed descriptively in the full analysis set and immunogenicity data were analyzed in the per protocol immunogenicity population. SARS-CoV-2 Spike (S)-binding antibody titers expressed as ELISA Unit per milliliters (EU/mL), and neutralizing antibody titers in the wtVNA, expressed as the reciprocal serum dilution neutralizing 50% of the test virus dose (50% inhibitory concentration [IC_50_]), are displayed on a log_10_ scale and described using geometric mean titers (GMT) and 95% confidence intervals (95% CIs). For both assays, seroconversion was defined as having an antibody titer above the lower limit of quantification (LLOQ) post vaccination if the baseline titer was below the LLOQ, or a 4-fold increase over baseline post vaccination if the baseline titer was above the LLOQ. ICS responses were described as percentage of total CD4+ or CD8+ T cell population. Sample positivity was determined with a one-sided Fisher’s exact test comparing non-stimulated versus S peptide stimulated wells. LLOQ was 0.022% and non-quantifiable values were imputed to LLOQ/2. Th1/Th2 ratio was calculated if the Th1 and/or Th2 responses were positive and above 2xLLOQ. If the Th1 or Th2 response from a participant was not fulfilling these criteria, the Th1/Th2 ratio was considered ≥1 if a Th2 response could not be measured and <1 if a Th1 response could not be measured.

## RESULTS

### STUDY PARTICIPANTS

Screening of participants started July 13, 2020, first vaccination of cohort 1a participants (age 18– 55) was initiated on July 22, 2020 and completed on August 4, 2020, and in cohort 1b participants initiated on July 29, 2020 and completed on August 7, 2020. 575 volunteers were screened, of whom 380 were enrolled in cohort 1a (377 vaccinated) and 28 were screened and 25 enrolled in cohort 1b (25 vaccinated; Figure 1A). Seven (1.7%) participants were seropositive for SARS-CoV-2 at screening, seropositivity rate for Ad26 will be reported later. Vaccinations of cohort 3 participants (age ≥65) were initiated on August 3, 2020 and first vaccinations were completed on August 24, 2020. 660 volunteers were screened, of whom 405 were enrolled and 403 were vaccinated (at the time of database extract, safety and reactogenicity data was available for 394 vaccinated participants; Figure 1B). Immunogenicity results of only the first 15 participants of cohort 3 were available at time of this publication.

Baseline characteristics were broadly comparable across groups (Table 2).

**Table 2.** Baseline demographics.

|  | C1a | C1b | C3 |
| --- | --- | --- | --- |
| Characteristics | All Participants | All Participants | All Participants |
| N | 377 | 25 | 394 |
| Age (years) |  |  |  |
| Median | 34.00 | 42.00 | 69.00 |
| Range | (18.0; 55.0) | (22.0; 52.0) | (65.0; 88.0) |
| Sex |  |  |  |
| Female (%) | 198 (52.5%) | 13 (52.0%) | 199 (50.5%) |
| Male (%) | 179 (47.5%) | 11 (44.0%) | 195 (49.5%) |
| Undifferentiated | 0 | 1 (4.0%) | 0 |
| Race |  |  |  |
| White | 342 (90.7%) | 22 (88.0%) | 385 (97.7%) |
| Black or African American | 20 (5.3%) | 0 | 3 (0.8%) |
| Asian | 8 (2.1%) | 2 (8.0%) | 0 |
| Native Hawaiian or other Pacific Islander | 1 (0.3%) | 0 | 0 |
| American Indian or Alaska Native | 3 (0.8%) | 0 | 1 (0.3%) |
| Multiple | 0 | 1 (4.0%) | 0 |
| Unknown | 3 (0.8%) | 0 | 1 (0.3%) |
| Not reported | 0 | 0 | 4 (1.0%) |
| Ethnicity |  |  |  |
| Hispanic or Latino | 15 (4.0%) | 2 (8.0%) | 6 (1.5%) |
| Not Hispanic or Latino | 358 (95.0%) | 22 (88.0%) | 385 (97.7%) |
| Unknown | 1 (0.3%) | 0 | 0 |
| Not reported | 3 (0.8%) | 1 (4.0%) | 3 (0.8%) |
| BMI (kg/m2) |  |  |  |
| Mean (SD) | 24.596 (3.1942) | 24.456 (3.2223) | 25.357 (2.8594)* |
| Range | (16.80; 29.90) | (18.80; 29.90) | (16.60; 29.90)* |
| SARS-CoV-2 Seropositivity status at baseline |  |  |  |
| Positive | 7 (1.9%) | 0 | 3 (0.8%) |

\*N=393

### VACCINE SAFETY AND REACTOGENICITY OF Ad26.COV2.S

Blinded safety data from cohort 1a, 1b and 3 were collected and analyzed. Solicited AEs were collected on diary cards for 7 days post vaccination, unsolicited AEs for 28 days after vaccination and SAEs throughout the course of the study. The study is ongoing, and the second immunization of the primary regimen will not be completed at the time of this submission. To ensure the participants and the investigators evaluating the participants remain blinded in this study until the last dose of the primary vaccination regimen is given, the safety data are presented without group unblinding in this interim report.

In cohorts 1a and 1b the investigator’s assessment of reactogenicity is available for 402 participants, of whom 288 (72%) participants have reported solicited AEs (Table 3). Solicited local AEs were reported in 235 (58%) participants – mostly grade 1/grade 2. Three participants reported grade 3 pain/tenderness. The most frequent AE was injection site pain. Solicited systemic AEs were reported for 258 (64%) participants, mostly grade 1/grade 2, with grade 3 systemic AEs reported for 46 (11%) participants. The most frequent AEs were fatigue, headache and myalgia. Fever was reported in 76 (19%) participants, with grade 3 fever reported in 22 (5%) participants. All fevers occurred within 2 days of immunization and resolved within 1 to 2 days. Of the participants who experienced grade 1 or 2 fevers, 83% utilized antipyretics at the onset of symptoms. 91% of the participants who experienced grade 3 fevers utilized antipyretics at the onset of symptoms. Overall, 98 participants have reported unsolicited AEs with 12 reporting grade 3 AEs (Table 4).

**Table 3.**
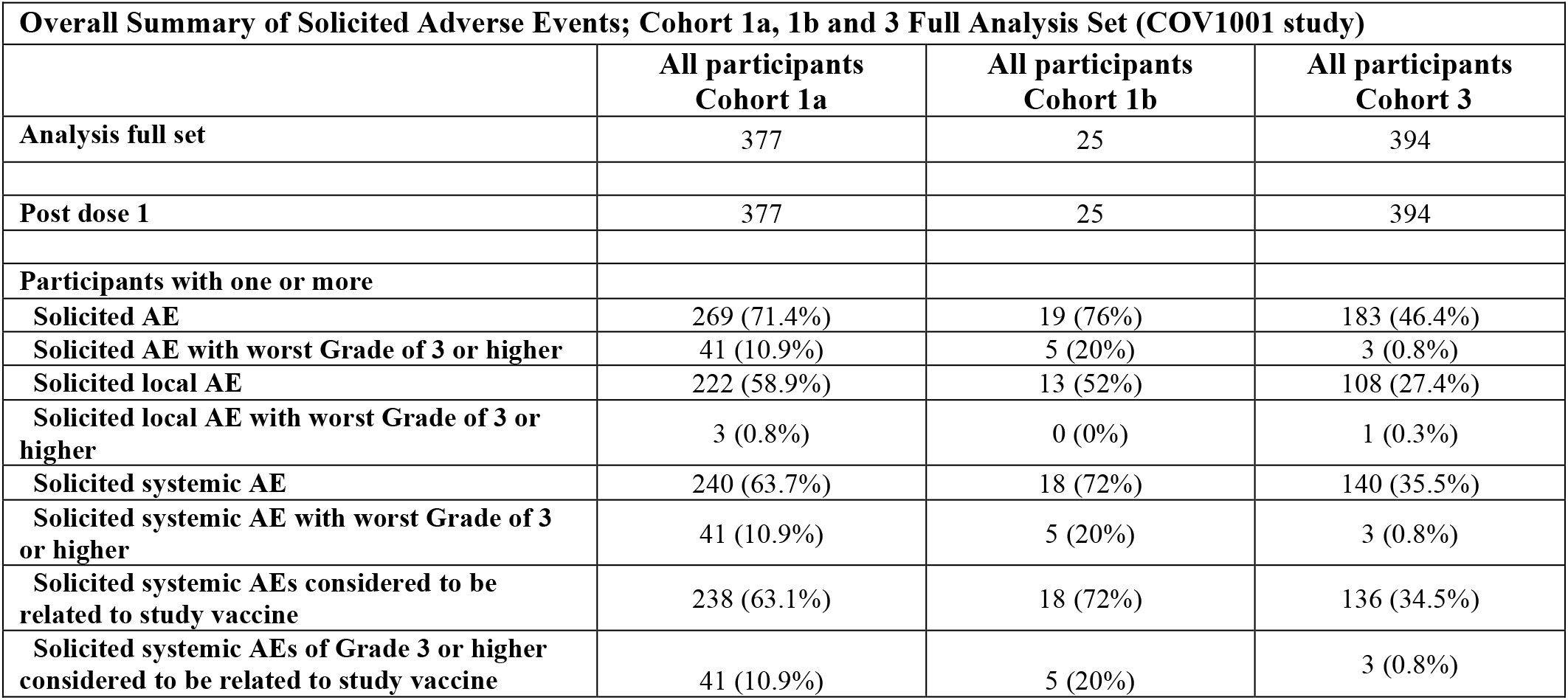
Summary of solicited AEs in all cohorts

| **Overall Summary of Solicited Adverse Events; Cohort 1a, 1b and 3 Full Analysis Set (COV1001 study)** |  |  |  |
| --- | --- | --- | --- |
|  | **All participants**<br>**Cohort 1a** | **All participants**<br>**Cohort 1b** | **All participants**<br>**Cohort 3** |
| **Analysis full set** | 377 | 25 | 394 |
| **Post dose 1** | 377 | 25 | 394 |
| **Participants with one or more** |  |  |  |
| **Solicited AE** | 269 (71.4%) | 19 (76%) | 183 (46.4%) |
| **Solicited AE with worst Grade of 3 or higher** | 41 (10.9%) | 5 (20%) | 3 (0.8%) |
| **Solicited local AE** | 222 (58.9%) | 13 (52%) | 108 (27.4%) |
| **Solicited local AE with worst Grade of 3 or higher** | 3 (0.8%) | 0 (0%) | 1 (0.3%) |
| **Solicited systemic AE** | 240 (63.7%) | 18 (72%) | 140 (35.5%) |
| **Solicited systemic AE with worst Grade of 3 or higher** | 41 (10.9%) | 5 (20%) | 3 (0.8%) |
| **Solicited systemic AEs considered to be related to study vaccine** | 238 (63.1%) | 18 (72%) | 136 (34.5%) |
| **Solicited systemic AEs of Grade 3 or higher considered to be related to study vaccine** | 41 (10.9%) | 5 (20%) | 3 (0.8%) |

**Table 4.** Grade 3 unsolicited AEs in Cohort 1a and cohort 3.

| Cohort | Age range | Sex | Preferred Term | Reported Term for the Adverse Event | Study Day of Start of Adverse Event | Study Day of End of Adverse Event | Serious Event | Causality | Action Taken with Study Treatment | Outcome of Adverse Event | Concomitant or Additional Tx Given |
| --- | --- | --- | --- | --- | --- | --- | --- | --- | --- | --- | --- |
| COHORT 1A | 40-50 | F | Blood pressure decreased | DECREASED BLOOD PRESSURE | 1 | 4 | Y | NOT RELATED | DOSE NOT CHANGED | RECOVERED/RESOLVED | Y |
| COHORT 1A | 50-55 | M | White blood cell count increased | WBC INCREASE | 8 | 12 | N | RELATED | DOSE NOT CHANGED | RECOVERED/RESOLVED | N |
| COHORT 1A | 18-25 | F | Malaise | MALAISE | 1 | 2 | N | RELATED | DOSE NOT CHANGED | RECOVERED/RESOLVED | N |
| COHORT 1A | 18-25 | F | Back pain | BACKPAIN | 2 | 2 | N | RELATED | DOSE NOT CHANGED | RECOVERED/RESOLVED | Y |
| COHORT 1A | 35-40 | F | Hypotensive crisis | HYPOTENSIVE CRISIS | 1 | 1 | N | RELATED | DOSE NOT CHANGED | RECOVERED/RESOLVED | Y |
| COHORT 1A | 30-35 | M | Insomnia | INSOMNIA | 2 | 2 | N | RELATED | DOSE NOT CHANGED | RECOVERED/RESOLVED | N |
| COHORT 1A | 18-25 | M | Contusion | CONTUSION RIGHT ANKLE | 5 |  | N | NOT RELATED | DOSE NOT CHANGED | RECOVERING/RESOLVING | Y |
| COHORT 1A | 30-35 | M | Pyrexia | FEVER | 1 | 2 | Y | RELATED | DRUG WITHDRAWN | RECOVERED/RESOLVED | Y |
| COHORT 1A | 25-30 | F | Back pain | BACKPAIN | 1 | 2 | N | RELATED | DOSE NOT CHANGED | RECOVERED/RESOLVED | Y |
| COHORT 1A | 18-25 | M | Dizziness | LIGHTHEADEDNESS | 2 | 2 | N | RELATED | DOSE NOT CHANGED | RECOVERED/RESOLVED | N |
| COHORT 1A | 18-25 | F | Heat stroke | SUNSTROKE | 13 | 13 | N | NOT RELATED | DOSE NOT CHANGED | RECOVERED/RESOLVED | Y |
| COHORT 1A | 40-45 | F | Neck pain | NECK PAIN | 1 | 16 | N | NOT RELATED | DOSE NOT CHANGED | RECOVERED/RESOLVED | Y |
| COHORT 3 | 65-70 | F | Dizziness | DIZZINESS | 6 | 7 | N | RELATED | DOSE NOT CHANGED | RECOVERED/RESOLVED | N |
| COHORT 3 | 65-70 | F | Vomiting | VOMITING | 6 | 7 | N | RELATED | DOSE NOT CHANGED | RECOVERED/RESOLVED | N |
| COHORT 3 | 75-80 | M | Systolic hypertension | GRADE 3 SYSTOLIC HYPERTENSION | 15 |  | N | NOT RELATED | DOSE NOT CHANGED | RECOVERING/RESOLVING | N |
| COHORT 3 | 70-75 | M | Hypertension | HYPERTENSION WORSENING | 8 |  | N | RELATED | DOSE NOT CHANGED | RECOVERING/RESOLVING | N |
| COHORT 3 | 65-70 | F | Systolic hypertension | SYSTOLIC HYPERTENSION (GRADE 3) | 1 | 1 | N | RELATED | DOSE NOT CHANGED | RECOVERED/RESOLVED | N |
| COHORT 3 | 65-70 | F | Bradycardia | WORSENING OF BRADYCARDIA | 1 | 1 | N | NOT RELATED | DOSE NOT CHANGED | RECOVERED/RESOLVED | N |

In cohort 3, at the time of database extract, investigator’s assessment of reactogenicity is available for 394 participants, of whom 183 (46%) participants have reported solicited AEs (Table 3). Solicited local AEs were reported in 108 (27%) participants, most of which were grade 1/Grade 2, with one participant reporting grade 3 swelling and erythema. The most frequent AE was injection site pain. Solicited systemic AEs were reported in 140 (36%) participants, most of which were grade 1/grade 2, with three participants reporting grade 3 AEs. The most frequent AEs were headache, fatigue and myalgia. Mild or moderate fevers of grade 1 or 2 were reported in 14 (4%) of participants only one of which was a grade 2. There were no high or other fevers that restricted daily living activities (grade 3) reported in cohort 3. Overall, 46 (12%) participants have reported unsolicited AEs. Four (1%) participants have reported unsolicited grade 3 AEs.

No grade 4 AEs, solicited or unsolicited, were reported in any cohort.

No participant discontinued the study due to an AE. There were two SAEs: one hypotension judged by the investigator to not be vaccine related because of a past history of recurrent hypotension, and one participant with fever who was hospitalized overnight because of suspicion of COVID-19, wo recovered within 12 hours, the fever was subsequently judged by the investigator to be vaccine related. For details see supplementary material.

While reactogenicity was acceptable in all groups, there was a trend for higher reactogenicity with the higher vaccine dose (data not shown) and with younger age. This will be reported in more detail upon group unblinding of the data.

### IMMUNOGENICITY OF Ad26.COV2.S

Immunogenicity data for this interim analysis were unblinded by dose level. Antibodies against a stabilized SARS-CoV-2 full length Spike protein were measured by ELISA. In cohort 1a, 94% and 98% of the participants that received the 5×10^10^ and 1×10^11^ vp dose, respectively, at baseline had EU/mL GMTs to SARS-CoV-2 S protein below the LLOQ. By Day 29 after vaccination, GMTs had increased to respectively 528 (95% CI: 442; 630) and 695 (95% CI: 596; 810), with 99% seroconversion in each dose group (Figure 2A). Of the cohort 1a participants who were seropositive at baseline, 7 out of 8 and 2 out of 3 demonstrated the preset criterion of a 4-fold increase in binding antibody titer to be considered a vaccine responder, for the 5×10^10^ and 1×10^11^ vp dose level groups, respectively. The initial 15 enrolled participants in cohort 3 were seronegative at baseline and had seroconverted by Day 29 post vaccination with GMTs of 507 (95% CI: 181; 1418) and 248 (95% CI: 122; 506), for the 5×10^10^ and 1×10^11^ vp dose level group, respectively. GMTs of 899 were observed in the human convalescent serum (HCS) panel used in this study, with an overlap in the 95% CI of the GMT for both dose level in each cohort.

**Figure 2:**
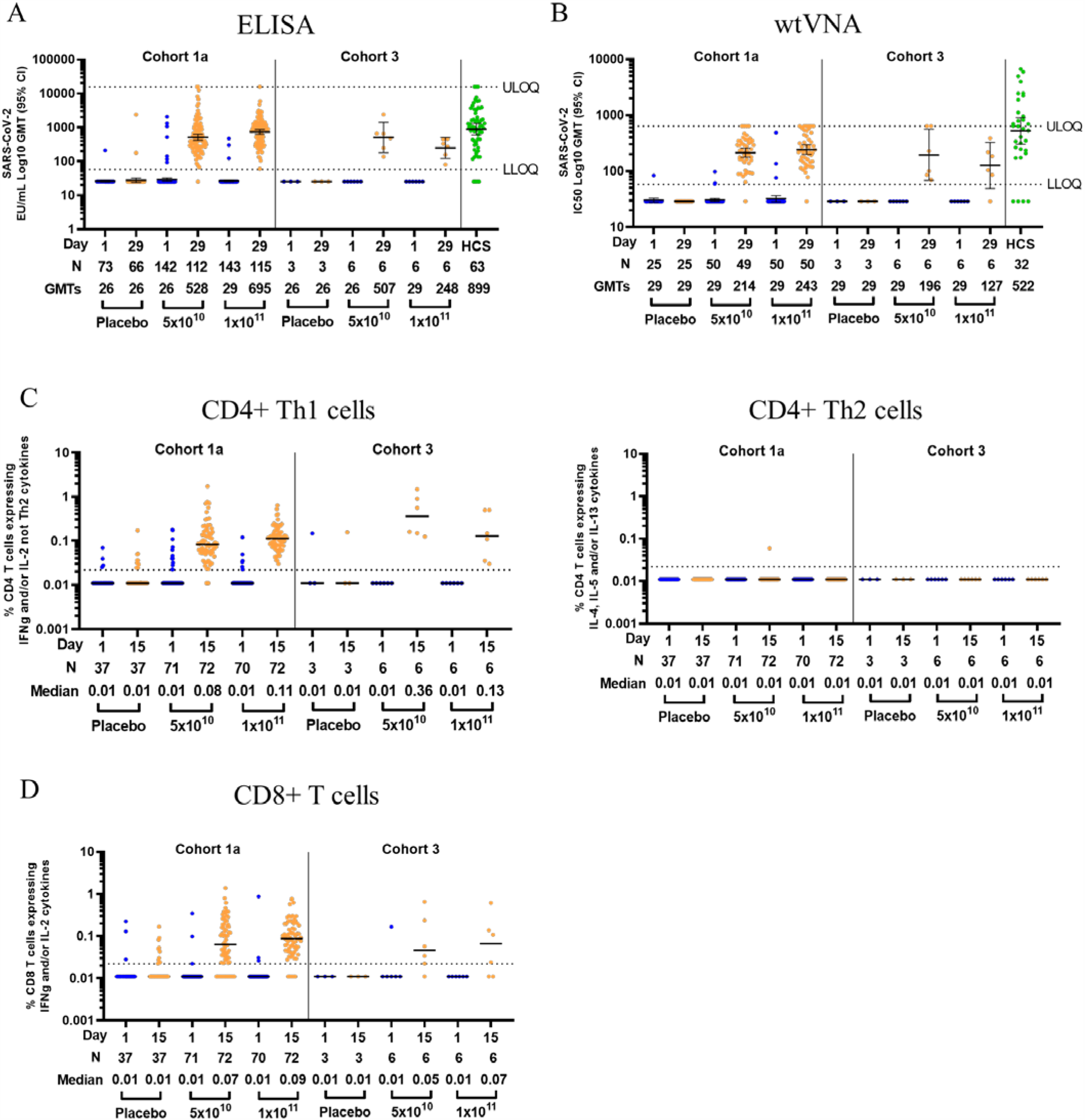
Immunogenicity of Ad26.COV2.S. (A) Log geometric mean titers (GMTs - as illustrated by the horizontal bars and the numbers below each timepoint) of SARS-CoV-2 binding antibodies in serum as measured by ELISA (ELISA Units per mL [EU/mL]), at baseline and at Day 29 post vaccination, among all participants, according to schedule in cohort 1a and 3. Dotted lines indicate the lower limit of quantification (LLOQ) and upper limit of quantification (ULOQ) of the assay, error bars indicate 95% confidence interval (CI). For values below the LLOQ, LLOQ/2 values were plotted. (B) Log GMTs of serum SARS-CoV-2 neutralizing antibodies, measured by 50% neutralization assay (IC_50_ Log GMT - as illustrated by the horizontal bars and the numbers below each timepoint), at baseline and at Day 29 post vaccination, among a subset of participants, according to schedule, in cohort 1a and 3. Dotted lines indicate the LLOQ and ULOQ of the assay run with the current pre-dilution used for vaccine samples, error bars indicate 95% CI. For values below the LLOQ, LLOQ/2 values were plotted. (C) Expression of Th1 (IFN-γ and/or IL-2, and not IL-4, IL-5 and IL-13), and Th2 (IL-4 and/or IL-5 and/or IL-13 and CD40L) cytokines by CD4+ T cells was measured by intracellular cytokine staining (ICS). Median (as illustrated by the horizontal bars and the numbers below each timepoint) and individual ICS responses to a SARS-CoV-2 S protein peptide pool in peripheral blood mononuclear cells, at baseline and 15 days post vaccination, among a subset of participants from cohort 1a and 3, according to schedule, are given. The Y-axis denotes the percentage of T cells positive for the Th1 or Th2 cytokines. Dotted line indicates the LLOQ. (D) Expression of IFN-γ and/or IL-2 cytokines by CD8+ T cells was measured by ICS. Median (as illustrated by the horizontal bars and the numbers below each timepoint) and individual ICS responses to SARS-CoV-2 S protein peptide pool in peripheral blood mononuclear cells, at baseline and 15 days post vaccination, among a subset of participants from cohort 1a and 3, according to schedule, are given. The Y-axis denotes the percentage of CD8+ T cells positive for IFN-γ and/or IL-2 cytokines. Dotted line indicates the LLOQ.

In a subset of participants (n=50) for each of the 5×10^10^ and 1×10^11^ vp dose level group, SARS-CoV-2 neutralizing antibody titers were measured by wtVNA. In cohort 1a, GMTs <LLOQ at baseline for both dose groups had increased to IC_50_ GMTs of 214 (95% CI: 177; 259) and 243 (95% CI: 200; 295) for the 5×10^10^ and 1×10^11^ vp group, respectively, by Day 29. Similar results were observed in the first 15 participants of cohort 3, with a GMT <LLOQ at baseline that had increased to GMTs of 196 (95% CI: 69; 560) and 127 (95% CI: <LLOQ; 327) for the 5×10^10^ and 1×10^11^ vp group, respectively, by Day 29 (Figure 2B). For comparison, a GMT of 522 was measured in the HCS panel used in this analysis, with an overlap in the 95% CI of the GMT between HCS panel and both dose groups and cohorts. Several serum samples from trial participants reached the upper limit of quantification (ULOQ = 640) for the vaccine sample analysis run and are currently being reanalyzed at higher dilution, which will raise the overall GMT for each dose level. At Day 29 post vaccination, 98% (97/99) of participants in cohort 1a were positive for neutralizing antibodies against SARS-CoV-2, independent of vaccine dose level that was given. The seroconversion rate at day 29 was 92% for both the 5×10^10^ and 1×10^11^ vp group in cohort 1a and, 100% (6/6) and 83% (5/6) for the 5×10^10^ and 1×10^11^ vp group in cohort 3. Importantly, in cohort 1a, a total of 82% and 94% of participants in the 5×10^10^ and 1×10^11^ vp group, respectively, reached titers greater than 100, indicative of a robust response induced in the vast majority of the participants after a single vaccination with Ad26.COV2.S. The wtVNA and ELISA titers as measured in cohort 1a samples highly correlated, with a Spearman correlation of 0.86 (p<0.001, data not shown). Data from a pseudovirus expressing SARS-CoV-2 S protein neutralization assay are pending and will be included in the full publication of our post dose 1 interim analysis.

SARS-CoV-2 S-specific CD4+ and CD8+ T cell responses were characterized in a subset of study participants at baseline and 15 days post vaccination. Previous studies with SARS-CoV and MERS-CoV vaccines in preclinical models have suggested an association between a Th2 skewed CD4+ T cell response with vaccine-associated enhanced respiratory disease (VAERD).^13–15^ To explore this theoretical risk of VAERD here, we assessed CD4+ Th1 and Th2 responses induced by Ad26.COV2.S. Following stimulation with peptides covering the whole S protein, median CD4+ Th1 responses increased from undetectable at baseline to a median of 0.08% (95% CI: 0.05; 0.16) and 0.11% (95% CI: 0.07; 0.16) 15 days post vaccination for the 5×10^10^ and 1×10^11^ vp group, respectively, in cohort 1a participants, and from non-detectable at baseline to a median of 0.36% (95% CI: 0.15; 0.89) and 0.13% (95% CI: 0.04; 0.50), respectively, in the first participants of cohort 3 (Figure 2C). In cohort 1a, 76% (95% CI: 65; 86) and 83% (95% CI: 73; 91) of participants had detectable Th1 responses for recipients of the 5×10^10^ and 1×10^11^ vp dose levels, respectively, and in the first 15 participants of cohort 3, 100% (95% CI: 54; 100) and 67% (95% CI: 22; 96), respectively. Only one participant in the 5×10^10^ vp group in cohort 1a had a detectable Th2 response. However, the Th1/Th2 ratio for this participant was 28.9, indicative of a Th1-skewed phenotype. In all other participants that had measurable Th1 and/or Th2 responses, the Th1/Th2 ratio ranged from 1.0 to 68.5. Overall, these data indicate that Ad26.COV2.S induced Th1-skewed responses in both age groups, suggestive for a low risk of VAERD.

S-specific CD8+ T cell responses were identified by the expression of IFN-γ and/or IL-2 cytokines upon S peptide stimulation (Figure 2D). 15 days post vaccination, the median magnitude of S-specific CD8+ T cell responses was 0.07% (95% CI: 0.03; 0.19) and 0.09% (95% CI: 0.05; 0.19) for the 5×10^10^ and 1×10^11^ vp group, respectively, in cohort 1a, and 0.05% (95% CI: 0.02; 0.24) and 0.07% (95% CI:0.02; 0.14), respectively, in cohort 3. 51% (95% CI: 39; 63) and 64% (95% CI: 52; 75) of participants in cohort 1a had a positive CD8+ T cell response to S peptide stimulation for the 5×10^10^ and 1×10^11^ vp group, respectively and 33% (95% CI: 4%; 78%) of participants of both dose level groups in cohort 3 had a detectable vaccine induced CD8+ T cell response.

## DISCUSSION

The interim analysis of our Phase 1/2a study shows that our vaccine candidate Ad26.COV2.S has an acceptable safety and reactogenicity profile and is immunogenic at both a 5×10^10^ or 1×10^11^ vp. Although the safety data in this interim report remain blinded, the overall occurrence independent of dose level of solicited systemic AEs of 64% with a 19% fever rate (5% grade 3) in adults aged 18 to 55 (Cohort 1a and 1b) stands in contrast to the solicited systemic AEs of 36% with a 4% fever rate (0% grade 3), found in the participants ≥ 65 years of age. This finding suggests that the vaccine candidate is less reactogenic in older adults. The safety profile is acceptable at any age given the seriousness of the disease the vaccine can potentially protect against and the nature of the pandemic, especially in the elderly which is the most vulnerable population to COVID-19. We did observe a trend for higher reactogenicity with the higher vaccine dose. More details on the safety and reactogenicity profile will be provided immediately upon group unblinding of the study, after vaccine dosing in cohort 1a and 3 has been completed.

A single dose of Ad26.COV2.S elicited strong humoral responses in the vast majority of vaccine recipients. S-binding antibody titers as measured by ELISA, increased from baseline to Day 29 post vaccination in 99% of the participants in cohort 1a and 100% of the first participants in cohort 3, independent of the vaccine dose level that was given. Similarly, high response rates were observed in a wtVNA. 29 days post vaccination, 98% of the participants had detectable neutralizing antibodies. 92% of cohort 1a participants and respectively 6 out of 6, and 5 out of 6 recipients of the 5×10^10^ vp and 1×10^11^ vp dose level in cohort 3, seroconverted for SARS-CoV-2 neutralizing antibodies. In cohort 1a, depending on the dose level, 82% to 94% of the participants had a neutralizing antibody titer above 100.

The lack of standards and the use of different immune assays by different vaccine developers makes it difficult to compare the performance of COVID-19 vaccine candidates that are currently in development. In addition, the level of immune response required to confer protection is unknown and even the least immunogenic vaccine may still elicit sufficient immunity to be protective. All other COVID-19 vaccines currently in development require two doses while neutralizing antibody responses in all participants reported here were obtained after a single dose of Ad26.COV2.S. The potency of a single vaccination with our Ad26.COV2.S COVID-19 vaccine candidate is supported by our recently reported study in non-human primates where a single dose provided complete protection against SARS-CoV-2 replication in the lung and near complete protection against viral replication in the nose.^5^ In this prior preclinical study, all sham vaccinated control animals had detectable virus in both lung and nose for 7–14 days. In vaccinated non-human primates, protection against SARS-CoV-2 infection was correlated with neutralizing antibody titers.

Our clinical data indicate that Ad26.COV2.S at both dose levels induces a strong neutralizing antibody response in the vast majority of the healthy young adult and older participants. Although only a limited number of participants of cohort 3 were included in the analysis so far, comparable immunogenicity of Ad26.COV2.S in adults aged 18–55 and adults aged 65–75 as observed here is encouraging for elderly individuals, who are at higher risk for developing severe COVID-19. Compared to COVID-19 human convalescent sera, the levels of binding and neutralizing antibodies induced by Ad26.COV2.S are in the same range for most participants. However, the significance of this comparison is not yet established due to the variability in GMTs in different human convalescent serum panels, likely related to the variability of the composition of these panels. Demographics such as age, disease severity, and time of sampling since disease onset and the number of samples in the panel could have an impact on the GMTs of antibody, making a comparison to GMTs in samples from vaccinated participants arbitrary.^16^

Previous experience with certain animal models of SARS-CoV and MERS-CoV vaccines have raised a theoretical concern of VAERD.^13–15^ An association between this safety concern and poor neutralizing potency of humoral immunity and Th2-skewed cellular immune responses has been suggested. Here we show that all Ad26.COV2.S elicited CD4+ T cell responses were Th1 skewed with no or very low Th2 responses. Indeed, in all responders, the Th1/Th2 ratio was above 1, in line with previous experience with our Ad26-based vaccine platform (data on file).^10^ This robust CD4+ Th1 response was accompanied by strong CD8+ T cell responses following vaccination. Robust CD4+ Th1 and CD8+ T cell responses, in addition to strong humoral responses elicited by Ad26.COV2.S minimizes the theoretical risk of VAERD.

An efficacious single-dose COVID-19 vaccine would have advantages over a two-dose vaccine in terms of implementation, especially during a pandemic. If a single dose of Ad26.COV2.S protects against SARS.CoV.2 infection or COVID-19, the durability of protection will be important to characterize. Data on the durability of the immune response elicited by a single dose of Ad26.COV2.S as well as on the immune response after a second dose of Ad26.COV2.S will become available from our ongoing Phase 1/2a studies (Table 1). In previous studies with the Ad26-based Zika vaccine, durability of neutralizing antibodies after a single dose, defined as seropositivity, was at least 12 months in 56% of vaccine recipients and a second dose further increased this percentage.^10^ Obviously, durability of immunity is not only reflected in the maintenance of neutralizing antibody titers but also in the quality of vaccine priming of the immune system resulting in immune memory. To this end, we will study the ability of vaccinated participants in our ongoing Phase 2a study (COV2001) to mount an anamnestic response to a very low dose of vaccine as a surrogate for exposure to SARS-CoV-2.

The interpretation of the interim analysis presented here for immunogenicity is limited by the relatively small sample size in cohort 3, the short follow-up, and the demographic characteristics of the study population, which may limit the generalizability of our findings. However, additional data will become available soon.

In conclusion, our interim analysis indicates that a single dose of Ad26.COV2.S, either 5×10^10^ vp or 1×10^11^ vp, is safe, well tolerated and highly immunogenic. Based on similar immunogenicity of both dose levels, we have selected the lower dose for further clinical evaluation. In our first Phase 3 study that has been initiated in the US, and that will later include sites in Africa, and South and Central America (Trial Number: NCT04505722), we will evaluate the efficacy of a single dose of 5×10^10^ vp of the Ad26.COV2.S vaccine candidate. An additional Phase 3 study is planned to assess the efficacy and durability of immunity of a two-dose schedule with 5×10^10^ vp of the Ad26.COV2.S vaccine.

## Data Availability

The data referred to in the manuscript will be available.

## ACKNOWLEDGEMENTS

We thank the clinical trial participants. We thank the entire COV1001 study team including the clinical sites. From Janssen Vaccines & Prevention, we thank our clinical statistical team, the clinical operations team, medical writers, and clinical immunology department for their assistance. The content is solely the responsibility and opinions of the authors.

## FUNDING SOURCE

This project was sponsored by Johnson and Johnson and funded, in part, by the Department of Health and Human Services Biomedical Advanced Research and Development Authority under contract HHS0100201700018C.

## AUTHOR CONTRIBUTIONS

The clinical protocol was developed by JVP. Investigators at the clinical sites collected clinical data. Immunogenicity testing for interim analysis was performed at Fred Hutch Cancer Research Center, USA, Public Health England, UK and Nexelis, Canada. MLG, JS, JH, DH, CT, GS, DB, FS, MD, JVH and HS were involved in data analysis and interpretation. All authors jointly wrote the manuscript, made the decision to submit the manuscript for publication, attest to the integrity of the study, the completeness and accuracy of the interim data, and the fidelity of the study to the protocol.

## DISCLOSURES

Janssen Pharmaceuticals and the US Army have a cooperative research agreement (Cooperative Research and Development Agreement, CRADA). Under the terms of the agreement, Janssen may provide financial resources to support work; there is no personal financial benefit from the arrangement. JS, MLG, GS, DH, CT, MG, JS, ST, EC, GS, JH, FS, MD, JVH and HS are all employees of Janssen Pharmaceuticals and may be Johnson & Johnson stockholders..

## Notes

### Clinical Trial

NCT04436276

### Author Declarations

The study was reviewed and approved by local ethics committees (Comite d Ethique Hospitalo-Facultaire Sain-Luc, Universite Catholique de Louvain on July 16, 2020) and institutional review boards (IRB) (approval by Advarra IRB on June 29 and July 10, 2020, for New Orleans Center for Clinical Research and Optimal Research sites, respectively)

## REFERENCES

1. WHO. Disease outbreak news. 2020. Emergencies preparedness, response. Pneumonia of unknown origin – China.https://www.who.int/csr/don/05-january-2020-pneumonia-of-unkown-cause-china/en/ 5 January, Accessed 12 Jan 2020.

2. Zhu N, Zhang D, Wang W, et al. A Novel Coronavirus from Patients with Pneumonia in China, 2019. N Engl J Med 2020;382:727–733.

3. Folegatti PM, Ewer KJ, Aley PK, et al. Safety and immunogenicity of the ChAdOx1 nCoV-19 vaccine against SARS-CoV-2: a preliminary report of a phase 1/2, single-blind, randomised controlled trial. Lancet 2020; 396:467–478. DOI: 10.1016/S0140-6736(20)31604-4.

4. Jackson LA, Anderson EJ, Rouphael NG, et al. An mRNA Vaccine against SARS-CoV-2 — Preliminary Report. New Engl J Med 2020; DOI: 10.1056/NEJMoa2022483. Online ahead of print. DOI: 10.1056/NEJMoa2022483.

5. Mercado NB, Zahn R, Wegmann F, et al. Single-shot Ad26 vaccine protects against SARS-CoV-2 in rhesus macaques. Nature 2020 DOI: 10.1038/s41586-020-2607-z. Online ahead of print. DOI: 10.1038/s41586-020-2607-z

6. Mulligan MJ, Lyke KE, Kitchin N, et al. Phase 1/2 study of COVID-19 RNA vaccine BNT162b1 in adults. Nature 2020; DOI: 10.1038/s41586-020-2639-4. Online ahead of print.

7. Bos R, Rutten L, JEM van der Lubbe, et al. Ad26-vector based COVID-19 vaccine encoding a prefusion stabilized SARS-CoV-2 Spike immunogen induces potent humoral and cellular immune responses. https://biorxiv.org/content/10.1101/2020.07.30.227470v1.

8. Anywaine Z, Whitworth H, Kaleebu P, et al. Safety and Immunogenicity of a 2-Dose Heterologous Vaccination Regimen With Ad26.ZEBOV and MVA-BN-Filo Ebola Vaccines: 12-Month Data From a Phase 1 Randomized Clinical Trial in Uganda and Tanzania. J Infect Dis 2019; 220:4–56. DOI: 10.1093/infdis/jiz070

9. Williams K, Bastian AR, Feldman RA, et al. Phase 1 Safety and Immunogenicity Study of a Respiratory Syncytial Virus Vaccine With an Adenovirus 26 Vector Encoding Prefusion F (Ad26.RSV.preF) in Adults Aged ≥60 Years. J Infect Dis 2020; 222:979– 988. DOI: 10.1093/infdis/jiaa193

10. Barouch DH, Tomaka FL, Wegmann F, et al. Evaluation of a mosaic HIV-1 vaccine in a multicentre, randomised, double-blind, placebo-controlled, phase 1/2a clinical trial (APPROACH) and in rhesus monkeys (NHP 13-19). Lancet 2018;392:232–243. DOI: 10.1016/S0140-6736(18)31364-3

11. Tostanoski LH, Wegmann F, Martinot AL, et al. Ad26 vaccine protects against SARS-CoV-2 severe clinical disease in hamsters. Nat Med 2020;DOI: 10.1038/s41591-020-1070-6. Online ahead of print.

12. Custers J., Kim D., Leyssen M., et al; Vaccines based on replication incompetent Ad26 viral vectors: Standardized template with key considerations for a risk/benefit assessment. Vaccine, in press

13. Deming D, Sheahan T, Heise M, et al. Vaccine Efficacy in Senescent Mice Challenged with Recombinant SARS-CoV Bearing Epidemic and Zoonotic Spike Variants. PLoS Med 2018;DOI: 10.1371/journal.pmed.0030525

14. Iwata-Yoshkawa N, Uda A, Suzuki T, et al. Effects of Toll-Like Receptor Stimulation on Eosinophilic Infiltration in Lungs of BALB/c Mice Immunized with UV-Inactivated Severe Acute Respiratory Syndrome-Related Coronavirus Vaccine. J Virol 2014;88:8597–8614. DOI: 10.1128/JVI.00983-14

15. Yasui F, Kai C, Kitabatake M, et al. Prior Immunization with Severe Acute Respiratory Syndrome (SARS)-Associated Coronavirus (SARS-CoV) Nucleocapsid Protein Causes Severe Pneumonia in Mice Infected with SARS-CoV. J Immunol 2008;181:6337–6348. DOI: 10.4049/jimmunol.181.9.6337 Moore JP, Klasse PJ. SARS-CoV-2 vaccines: ‘Warp Speed’ needs mind melds not warped minds. J Virol 2020;94:e01083–20. doi: 10.1128/JVI.01083-20

